# Childbirth experiences in low-income communities: A qualitative case study of mothers in rural western Uganda

**DOI:** 10.1101/2025.08.18.25333395

**Authors:** Jonathan Mwiindi, Rigoberto I. Delgado, Beth Wangigi, Priscilla Busingye, Margarita Delgado Thompson, Jeseena Jabbar

## Abstract

**Background:** This study analyzed the birth experiences of rural Ugandan mothers who delivered by emergency C-section, and who received free care sponsored by a not-for-profit organization. There is little research done on the birth experience in sub-Saharan Africa, particularly on childbirth experience during C-section. The results from this study should provide information to improve the quality of maternal services for low-income families in rural African communities.

**Methods:** The study methodology included in-depth, semi-structured, open-ended patient interviews with eight post-natal young mothers (ages 18-25) who received emergency C-section care sponsored by the SAFE program in Nyakibale and Rushoroza hospitals, Uganda. The interviews were recorded in two local languages, Runyankole and Rukiga, and later translated and transcribed into English. The sample was of convenience and included women who were para 2 after delivery and reported patient experience as measured by the Hospital Consumer Assessment of Healthcare Providers and Systems patient experience tool. The data analysis was based on a thematic review approach.

**Results:** Several study participants were delayed in securing appropriate emergency obstetric care due to inexperience with unexpected labor signs, long distance between home and delivery hospital, access to transportation. Most participants expressed a positive delivery experience during labor and postpartum. Although the sponsoring organization covered all surgery and medical bills, other costs, such as transportation and food were not included, creating distress among the participating mothers.

**Conclusion:** A positive birth experience improves the health of the mother and her child, the mother–child relationship, and overall family well-being. The challenges faced by low-income mothers in rural areas of Uganda, highlight the importance of expanding targeted childbirth preparation and education. The delivery experience can be greatly enhanced with strict quality of care protocols like those implemented by the SAFE program, which include creating a culture of respect for the mother. Direct and indirect costs of healthcare in Uganda are a big challenge for low-income mothers in rural areas who pay out of pocket for these services. Programs like SAFE mitigate economic hardships of poor families and can serve as financing models for government institutions or other NGOs.

## Introduction

The experience of labor and birth is defined as an individual’s life event, incorporating interrelated psychological and physiological processes influenced by social, environmental, organizational, and policy contexts [1–3]. The quality of this experience affects the health of the mother and her child, the mother–child relationship, and spouse [4]. Several studies have been done on birth experiences in low- and middle-income countries (LMICs), however, there has been less research on birth experience in sub-Saharan Africa [5]. Furthermore, there is even less evidence on childbirth experience for expensive emergency cases in poor communities. This study analyzed the birth experience of rural Ugandan mothers who delivered by emergency C-section and participated in the Surgical Access for Everyone (SAFE) program; a funding initiative sponsored by African Mission Healthcare (AMH), a not-for-profit organization supporting low-income mothers with free maternal emergency care in Africa [6]. In addition, the SAFE program requires participating hospitals to implement strict measures of quality of care. The prevalence of emergency C-sections in sub-Saharan Africa is estimated at 4.6% [7]. Studying the experiences of SAFE participants allowed us to control an important factor in childbirth, costs of care.

Many factors play a role in defining positive and negative childbirth experience affecting the mother in different ways [8]. A positive childbirth experience, for example, is crucial in developing a woman’s psychological and emotional health throughout and following the perinatal period. These foster a mother’s self-confidence, feelings of accomplishment, better adjustment to motherhood, as well as improved bonding between mother and baby [9]. A positive birth experience is not only determined by clinical outcome, but also by following a woman-centered approach, which includes creating an environment where women feel respected, safe, and inclusive throughout the birthing process [10,11]. It is also correlated with the quality of communication offered by the healthcare provider [12,13].

Negative experiences are characterized by fear, excessive pain, poor support and care, discomfort, and undesirable outcomes affecting a woman’s emotional well-being and willingness to have another baby [14]. These experiences can result in postpartum depression and childbirth-related PTSD in mothers, increase risk of depression in partners, strained familial relationships, and can cause physiological or developmental abnormalities in children [15–17]. Studies show that up to 44% of women can describe childbirth experiences negatively, and 3% of male partners have indicated that their partner’s childbirth was a negative experience [8,18].

The results from this study should provide organizations like AMH with evidence need to improve the quality maternal and neonatal services for low-income families in rural African communities.

## Methods

The study methodology included in-depth, semi-structured, open-ended patient interviews with eight post-natal mothers who received emergency C-section (ECS) care sponsored by the SAFE program in Nyakibale and Rushoroza hospitals, Uganda. These hospitals are located in Southwestern Uganda, between 308 and 410 km from Kampala, the capital city and serve approximately 40,000 patients annually. Nyakibale has a bed capacity of 200 with 90 healthcare workers while Rushoroza is a 93-bed hospital with 34 health workers. The sample was of convenience and included women between the ages of 18 and 25 years of age, who were para 2 past delivery, and reported patient experience as measured using the Hospital Consumer Assessment of Healthcare Providers and Systems (HCAHPS) tool [19].

The interviews were part of the quarterly audit process conducted by the SAFE ECS Program and completed by a trained independent interviewer in two local languages, Runyankole and Rukiga. All interviews were conducted and audio recorded in April 2020. The de-identified transcripts were made available to the research team during the June - July 2020 period, after completing the translation into English, back-transcription, and verification for accuracy of all records. All data was de-identified for analysis purposes, and the research team had no access to information that could identify individual participants during or after data collection or analysis.

Data analysis was based on a thematic review approach as described by Braun and Clarke [20]. This included familiarization with transcripts of all the audio-recorded interviews and grouping responses by patient experience constructs as measured by the HCAHPS tool. Comparable results were matched into relevant sub-categories, which were merged into different themes. The study received approval from the Institutional Review Committee at the Mbarara University of Science and Technology, the Ugandan National Committee for Science and Technology, and from the Institutional Review Board at The University of Texas Health Science Center at Houston.

## Results

As shown in table 1, eight mothers participated in this study ranging in age between 20 and 25 years old. Their occupation varied between casual laborers and subsistence farming. The major indicators for emergency C-section among these women included previous scar, maternal compromise, fetal distress, and dystocia.

**Table 1:**
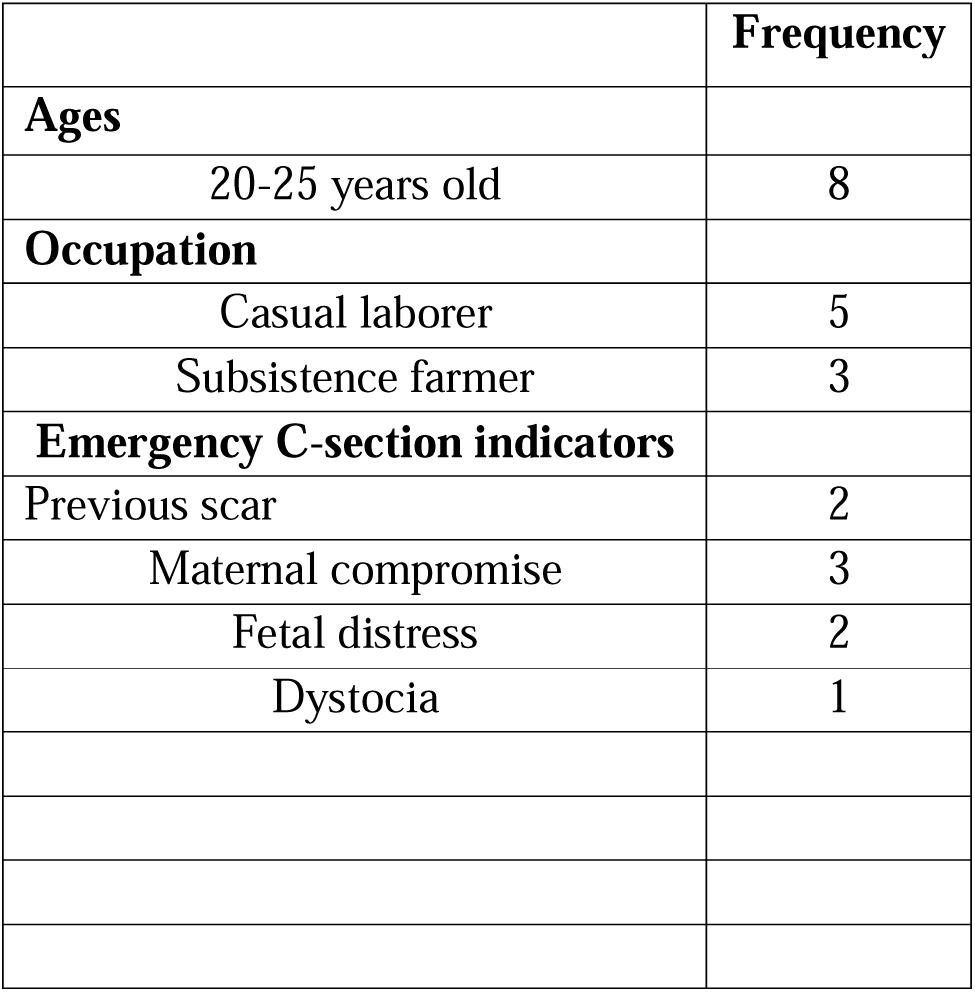
Demographic Characteristics of the Study Participants (n=8)

|  | Frequency |
| --- | --- |
| **Ages** |  |
| 20-25 years old | 8 |
| **Occupation** |  |
| Casual laborer | 5 |
| Subsistence farmer | 3 |
| **Emergency C-section indicators** |  |
| Previous scar | 2 |
| Maternal compromise | 3 |
| Fetal distress | 2 |
| Dystocia | 1 |

From the analysis of the interview transcripts, two major themes were established: 1) childbirth experience; and 2) post-operative financial concerns (Table 2). Three sub-themes emerged under childbirth experience, namely: early labor and decision to seek healthcare, experience at the health facility, and delivery experience. Typical issues within the theme on post-operative and financial concerns were additional costs of care while hospitalized. The following descriptions do not use the real names for mothers.

**Table 2:** Framework used to classify responses from each mother.

| Major Themes and Sub-themes | Typical Issues |
| --- | --- |
| ***Theme 1 - Childbirth experience*** |  |
| Early labor and decision to seek healthcare | Identifying correct symptoms; trusted family members influencing decisions to seek care. |
| Experience at the health facility | Degree of respect for patient defines quality of experience |
| Delivery experience |  |
| ***Theme 2 - Post-operative financial concerns*** | Covering food costs post-surgery and cost of care |

### Childbirth experience: Early labor and decision to seek care

The participants described early labor experiences.

> *"I did not feel it coming, but at around 11:00 p.m., my water broke, and I knew it was time” (Joan)*.

Half of the participants had visited health centers near their homes for antenatal care, and so they went to those health centers first when they needed to deliver. Joan and her husband walked 20 minutes to a health center close to their village, while Grace visited her parish health center first.

Not all participants were not able to go to hospital or health center immediately when they realized that they were in labor. Nancy, "started getting contractions at 8 p.m. but was not able to go to the hospital until morning," because she had neither the means nor the money for transport. Her husband had to borrow money from a neighbor to cover transportation costs to the hospital, until approximately 12 hours after onset of labor. This delay due to lack of transportation was not unique to Nancy. Other mothers were also affected, but partly due to a government-imposed COVID19 pandemic restrictions over vehicle or motorcycle circulation past 7p.m. One mother noted:

> *“My labor started at around 2.00 a.m. however, I had to wait until it was bright in the morning because during that time there was lockdown due to the Corona outbreak and it wasn’t easy to get means of transport in the middle of the night” (Rose)*.

Participants were later referred to higher-level care hospital facilities when staff at the health centers recognized that they needed C-sections. These initial clinics lacked experienced clinical staff and required equipment to perform surgical deliveries. In Joan’s case, the nurse at the health center determined that labor and delivery were too "complicated for her to manage," and was referred to Nyakibale hospital. For Rose, the health center nurse told her that they did not have the sutures required for the C-section. Grace was given a trial of labor at the health center for approximately 5 hours with attempts to increase contractions with medication. However, her labor didn’t progress well, became too weak and was referred to hospital by the health center nurse. She explained:

> *“…the contractions severely increased, and I became so weak. I couldn’t walk or even speak. So at around 3am that night I became so weak and could not even do anything for myself. At that time, she called my mum and told her that she had tried her best and due to the fact that I was getting very tired she wanted to refer me to another hospital because I could deliver when I am too tired for her to manage” (Grace)*.

The participants who had seen practitioners at hospitals for their antenatal care visits went straight to the hospital when they realized they were in labor.

While most mothers expressed confidence to cope with these symptoms of early labor, others were quite anxious; especially Ann who had initially missed signs of early pregnancy.

> *"I was given some dates, I think the eighth, but labor came on the fifth, and because I did not know about this pregnancy, I had miscalculated my days" (Ann)*.

She became concerned about a miscarrying when she realized that she had vaginal bleeding. Despite her husband and mother-in-law trying to convince her that she was not in labor yet, she insisted on going to hospital.

### Childbirth experience: Experience at the delivery hospital

Most of the participants expressed good interaction with the healthcare workers, at the health center where they first attended and at the delivery hospital. They defined this interaction as helpful and assuring. Those mothers who visited the health center first received clear explanations by the center staff on why they needed to be transferred to a hospital with critical care capacity.

Irene, who spent three days in Nyakibale hospital before being operated on described her interaction with the nurses:

> *“…they would explain to me all that was to be done. They would tell me their findings until they said labor was no longer progressing and that the baby was going to get tired … With the increasing contractions I was becoming weaker, and I accepted [to have] the operation” (Irene)*.

Participants felt nurses and doctors were informative and caring during their hospital stay. They felt cared for during their C-sections and post-op to determine pain medication as Sarah recounts:

> *“At first when I had just come out of theatre, I didn’t feel much pain but when it came to around 10pm, I felt a lot of pain, the area with the urinary catheter felt like burning fire and my abdomen was hurting. But the nurse who had taken me to the theatre happened to be passing by, when I told him, he rushed and brought me some medicine, an injection and the pain reduced. At around 3am this pain was back but the nurse had returned to check on me and he asked how I was feeling, I told him that I was feeling the same burning pain where the catheter was every time I would attempt to get out of bed. He told me I would be fine and added me more medicine and I became better; he really did everything for me.”*

There were exceptions, however. Rose, who was referred to hospital after failed attempts to augment labor in the health center, was upset that the hospital nurses did not understand the challenges that prevented her from coming to the hospital initially.

### Childbirth experience: Delivery experience

Most participants had anticipated that they would deliver vaginally but learned later they would be needing a C-section operation after arriving at the hospital. One important reason was a failed trial of labor and requiring surgical delivery. For example, Nancy:

> *“…when the baby wasn’t descending, the Nurse was there to counsel me, she assured me to wait and be patient. Even when I failed to progress, she told me that I had failed and explained to me that since I am not progressing, I will have to be taken for an operation.”*

One mother who had expected to deliver vaginally was too tired and weak by the time they reached Nyakibale hospital to have a good understanding of the pending operation. Before surgery was performed, however, she indicated she had recovered enough to provide consent.

Irene, who had also hoped to deliver vaginally, learned that a build-up of amniotic fluid around her baby could mean she would require a C-section.

These mothers faced a degree of anxiety, especially for those who did not know ahead of time that they required C-sections. They did not know what to expect and were afraid for their lives and the lives of their babies:

> *“I was worried about my life after the operation. I had never seen anyone they had operated on. I was also meditating on what I had been told that the baby might come out with the uterus or lose his life…” (Grace)*.

Other mothers overcame fear of surgery by reflecting on positive outcomes of the procedure:

> “*I also wished for the same thing, operation. I wanted the baby to come out alive but also for my life to be spared. I was really suffering. My life was at stake” (Joan)*.

Two mothers, Sarah and Hellen, knew they would need a C-section since both had required the procedure for their first child, and not enough time had passed to have a safe vaginal delivery with a second child. Sarah was only six months postpartum with her first child when she became pregnant with her second. She recounted:

> *“The man from the organization] asked me why I was operated on for my first delivery, and I told him that I did not have contractions, and the doctors told me that I had to deliver by an operation. I also told him that I got pregnant when my first child was still very young because she was only six months when I got pregnant.”*

Intraoperative experiences for most mothers also elicited feelings of fear and anxiety, and in some cases excessive pain:

> *“When in theatre I was received well, and I think they had prepared everything they needed. But when they started operating, I felt the cut. I don’t know what happened because when they made the cut, I responded then they asked me whether I had felt pain, and I said yes. So, they decided to put me to sleep completely, and I wasn’t aware of my surroundings during the operation” (Ann)*.

### Post-operation financial concerns

Although the sponsoring organization, SAFE, financed all surgery and medical bills, other costs incurred by the participating mothers, such as transportation and food, were not included. These mothers stayed in hospital between 3 or 4 days following their surgery, incurring costs of transportation to and from hospital, between health centers in early labor, and for caretakers to run errands while the participants were in the hospital. Hospitals in Uganda require patients to provide their own food during their stay, normally purchased at surrounding kiosks. Most participant mothers were told not to eat immediately following surgery, so food purchases were not imperative at first. However, when they could eat, their caretakers provided them with soft foods like rice and soup when available. During the study period options for buying food were limited due to COVID-19 lockdowns, which implied higher-cost meals for the mothers participating in the study.

Most of the participants said that without the SAFE funding, it would be very difficult for them to afford a hospitalization. Several mentioned that they would have needed to sell part of their land at ‘throw away’ prices or facing serious consequences for failing to pay costs of hospitalization: “*I think we would be jailed*” (Ann). One mother mentioned the choice of providing in-kind services to cover hospital costs:

> *“I don’t really know. Maybe I would have stayed and worked for the hospital because I wanted my life to be saved. I don’t think I had a way of raising this money” (Irene)*.

Several mothers accrued debt in the process and had to do some casual work. Ann, for example, mentioned having to “*work in someone’s garden for two days*” and earning ten thousand Uganda Shillings (2.65 USD; calculated at the exchange rate of USD 1= UGX 3,600). At the time of the interview, two months after her discharge from hospital, Ann and her husband were still in debt for all the money they had to borrow.

Some mothers indicated that the money spent on food while at the hospital was provided for by family members or savings. One mother said,

> *“I have a brother who sent me fifty thousand shillings (50,000/=), then my mother gave me thirty thousand shillings (30,000/=) then I also had twenty thousand shillings I had saved” (Irene)*.

All participating mothers expressed difficulties paying for additional food and transportation expenses, except for two who incurred extra cost of staying in private rooms at the hospital. This was because they had relatives that worked within the same facilities and offered to help with covering additional charges.

## Discussion

This study focused on understanding the birth experience of rural Ugandan mothers who deliver by emergency cesarean section and participated in a funding program covering costs of maternal emergency care for low-income families in Africa. The results show delays in securing appropriate emergency obstetric care due to inexperience with labor signs, access to transportation, and financial resources. These are similar factors found in other countries [21–23]. The reflections given by the study participants, however, highlight opportunities for clinicians, healthcare administrators and policy makers to introduce strategies for improving childbirth experience in low-income regions.

First, most mothers in the study knew that they were in labor after experiencing commonly known symptoms such as contractions. But other mothers had unexpected signs, creating anxiety and confusion. This highlights the importance of implementing childbirth education for pregnant mothers, particularly for dealing with issues of fear and pain [24] [25].

Ugandan cultural norms significantly impact a woman’s decision to seek health care. Specifically, decision-making regarding health care is not the woman’s jurisdiction alone, but rather the family as a unit [26–28]. In this study, most mothers considered their husband and/or mother-in-law as trusted decision-makers regarding when to go to a health facility, and which one to visit. This shows that to promote timely procurement of medical care it is important to include in the prenatal education sessions key members of the mother’s family.

Second, securing transportation, especially at night, was described as a major challenge for the study participants. In rural Uganda most women do not have the means or the money for transport. Not surprising, the decision of where to seek care for most mothers was based on proximity to their homes, as opposed to the quality of the services offered at a specific clinic. These facilities often lack the three elements defined as essential capacity to perform C-sections: 1) uninterrupted electricity, 2) staff (e.g., medical officer and anesthetist), and 3) availability of general anesthesia equipment and supplies [29,30]. Traveling to a well-resourced delivery hospital often requires women to either walk or use motor bicycles for more than two hours, which is seen as another leading cause of delays for treating complicated deliveries [31]. This finding is consistent with results from a study in Tanzania showing that common barriers for women seeking health services were lack of money for transport and long distances to health services[32–34]. Similarly, a study in Kenya found that government restrictions on travel and gatherings during the COVID season limited access to care and negatively impacting women’s health [35]. Thus, there is the need to explore the potential to expand remote medical support strategies such as telehealth or mobile, which have shown promise in several regions of Africa [36–38].

Third, regarding their stay at the hospital, most mothers reported good interaction with the hospital staff as they felt respected, mainly due to the patient care procedures implemented by the SAFE program. These are strict quality of care protocols which include four recognized factors which foster a positive facility-based childbirth experience: 1) responsiveness of the hospital staff; 2) supportive care from family, 3) dignified care where the women felt treated with dignity; and 4) effective communication from healthcare workers [39] [40,41].

Establishing a culture of respect for the mother is key for providing a positive birth experience and for creating trust in the community. For example, a study of primary care centers in Tanzania found that women who did not report disrespectful treatment were 2.1 times more likely to recommend the clinic to other women [42,43].

Fourth, although the SAFE program covered the costs of the emergency surgery, patients still needed to cover other costs, mainly food and laundry. Most Ugandan hospitals, especially those in rural areas, do not provide meals for patients. Participants in this study mentioned that it was difficult for them to raise money to buy meals during their hospital stay, requiring their caretakers to borrow money to cover additional costs. This finding, although not generalizable, is consistent with what other researchers have found showing that costs in public hospitals are significant hindrances to patients’ access to care [44][45]. All patients in this study could not afford to pay USD 250 (895,811 UGX) —the average C-section cost in Uganda. Paying such an amount would have devastated these families’ resources, and would have forced them to rely on third-party payers such as extended family members, government institutions, or donor programs like SAFE [46,47]. These results highlight the lack of any form of health insurance which forces families to meet all their medical expenses out-of-pocket.

This situation is not unique to the study participants. According to the World Bank, about 40% of total Ugandan health care expenditures are out-of-pocket, and nearly 80% of the population are at risk of catastrophic or impoverishing expenditures due to surgical care [48] [26]. Expanding donor programs like SAFE would greatly increase access to emergency care and reduce Uganda’s maternal mortality, which is one of the highest in Africa at 368 deaths per 100,000 [49] [50–52]. Some countries are implementing universal health coverage (UHC); however, success of these programs has been limited [53,54]. The typical UHC user fee exemption and waiver policies for the poor and vulnerable groups can potentially offer financial protection, but these policies are not widely available in many settings due to inadequate budgetary allocation to the health sector [55][56]. Another major obstacle for LMICs is funding; the overall costs of preventing maternal, fetal, and neonatal deaths alone is estimated at over US $4.5 billion [57]-[58,59].

## Conclusion

The experience of labor and birth impacts the health of the mother and her child, the mother–child relationship, and overall family well-being. The challenges faced by low-income mothers in rural areas of Uganda, highlight the importance of expanding targeted childbirth preparation and education. The delivery experience can be greatly enhanced with strict quality of care protocols like those implemented by the SAFE program, which include creating a culture of respect for the mother. Direct and indirect costs of healthcare in Uganda are a big challenge for low-income mothers in rural areas who pay out of pocket for these services. Programs like SAFE mitigate economic hardships of poor families and can serve as financing models for government institutions or other NGOs.

## Data Availability

The data underlying the results presented in the study are available from African Mission Healthcare. To request any data, please contact the manuscript's main author, Dr. Jonathan Mwiindi at. Note that only de-identified data can be shared upon request.

## Acknowledgments

The authors thank Dr. Joseph Kolars, Dr. Wendy Nyamusana, Sr. Gertrude Kabanyomozi, Sr. Miria Mazire, and Sr. Mary Mangdalene for their assistance in collecting data and revising the manuscript.

## Author Contributions

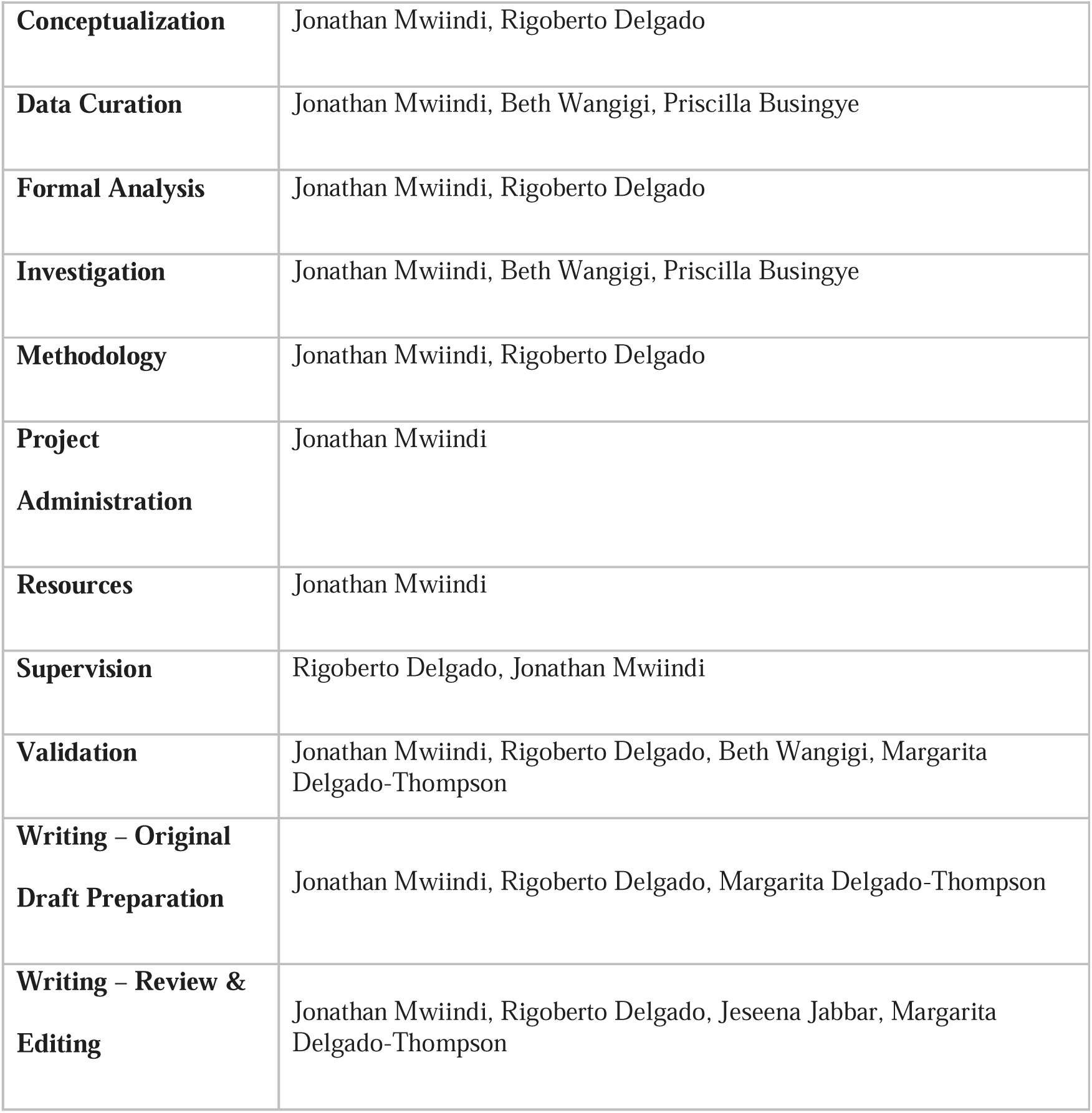

## Notes

### Competing Interest Statement

The authors have declared no competing interest.

### Funding Statement

The author(s) received no specific funding for this work.

### Author Declarations

Ethics committee/IRB of Mbarara University of Science and Technology gave ethical approval for this work. Ethics committee/IRB of The University of Texas Health Science Center at Houston gave ethical approval for this work. Ethics committee/IRB of the Ugandan National Committee for Science and Technology gave ethical approval for this work.

